# Protection against reinfection with SARS-CoV-2 omicron BA.2.75^*^ sublineage

**DOI:** 10.1101/2022.10.29.22281606

**Authors:** Hiam Chemaitelly, Patrick Tang, Peter Coyle, Hadi M. Yassine, Hebah A. Al-Khatib, Maria K. Smatti, Mohammad R. Hasan, Houssein H. Ayoub, Heba N. Altarawneh, Zaina Al-Kanaani, Einas Al-Kuwari, Andrew Jeremijenko, Anvar H. Kaleeckal, Ali N. Latif, Riyazuddin M. Shaik, Hanan F. Abdul-Rahim, Gheyath K. Nasrallah, Mohamed G. Al-Kuwari, Adeel A. Butt, Hamad E. Al-Romaihi, Mohamed H. Al-Thani, Abdullatif Al-Khal, Roberto Bertollini, Laith J. Abu-Raddad

## Abstract

The BA.2.75* sublineage of SARS-CoV-2 B.1.1.529 (omicron) variant escapes neutralizing antibodies. We estimated effectiveness of prior infection in preventing reinfection with BA.2.75* using a test-negative, case-control study design. Effectiveness of prior pre-omicron infection against BA.2.75* reinfection, irrespective of symptoms, was 6.0% (95% CI: 1.5-10.4%). Effectiveness of prior BA.1/BA.2 infection was 49.9% (95% CI: 47.6-52.1%) and of prior BA.4/BA.5 infection was 80.6% (95% CI: 71.2-87.0). Effectiveness of prior pre-omicron infection followed by BA.1/BA.2 infection against BA.2.75* reinfection was 56.4% (95% CI: 50.5-61.6). Effectiveness of prior pre-omicron infection followed by BA.4/BA.5 infection was 91.6% (95% CI: 65.1-98.0). Analyses stratified by time since prior infection indicated waning of protection since prior infection. Analyses stratified by vaccination status indicated that protection from prior infection is higher among those vaccinated, particularly among those who combined index-virus-type vaccination with a prior omicron infection. A combination of pre-omicron and omicron immunity is most protective against BA.2.75* reinfection. Viral immune evasion may have accelerated recently to overcome high immunity in the global population, thereby also accelerating waning of natural immunity.

The BA.2.75* sublineage of SARS-CoV-2 B.1.1.529 (omicron) variant escapes neutralizing antibodies.^1^ BA.2.75* (predominantly BA.2.75.2) became the dominant sublineage in Qatar by September 10, 2022 (Section S1 of the Supplementary Appendix and Figure S1). We estimated effectiveness of prior infection in preventing reinfection with BA.2.75* using a test-negative, case-control study design^2,3^ (Section S2).

We extracted data regarding SARS-CoV-2 laboratory testing, clinical infection, vaccination, and demographic details from the national SARS-CoV-2 databases, which include all results of polymerase chain reaction (PCR) and rapid antigen testing conducted at healthcare facilities in Qatar. Cases (SARS-CoV-2-positive tests) and controls (SARS-CoV-2-negative tests) were matched exactly according to sex, 10-year age group, nationality, number of coexisting conditions, number of vaccine doses at time of SARS-CoV-2 test, calendar week of testing, method of testing, and reason for testing, to control for differences in infection risk.^2^ Prior infections were classified as pre-omicron if they occurred before onset of the omicron wave on December 19, 2021 and as omicron otherwise.^3^ Omicron infections were classified by subvariant/sublineage according to when they dominated incidence: from December 19, 2021-June 7, 2022 for BA.1/BA.2,^3^ from June 8, 2022-September 9, 2022 for BA.4/BA.5,^3^ and from September 10, 2022-October 18, 2022 for BA.2.75*.

Figure S2 describes selection of the study population. Table S1 shows study population characteristics. Study population was broadly representative of Qatar’s population (Table S2). Effectiveness of prior pre-omicron infection against BA.2.75* reinfection, irrespective of symptoms, was 6.0% (95% CI: 1.5-10.4%; Figure 1A and Table S3). Effectiveness of prior BA.1/BA.2 infection was 49.9% (95% CI: 47.6-52.1%) and of prior BA.4/BA.5 infection was 80.6% (95% CI: 71.2-87.0). Effectiveness of prior pre-omicron infection followed by BA.1/BA.2 infection against BA.2.75* reinfection was 56.4% (95% CI: 50.5-61.6). Effectiveness of prior pre-omicron infection followed by BA.4/BA.5 infection was 91.6% (95% CI: 65.1-98.0).

**Figure 1.**
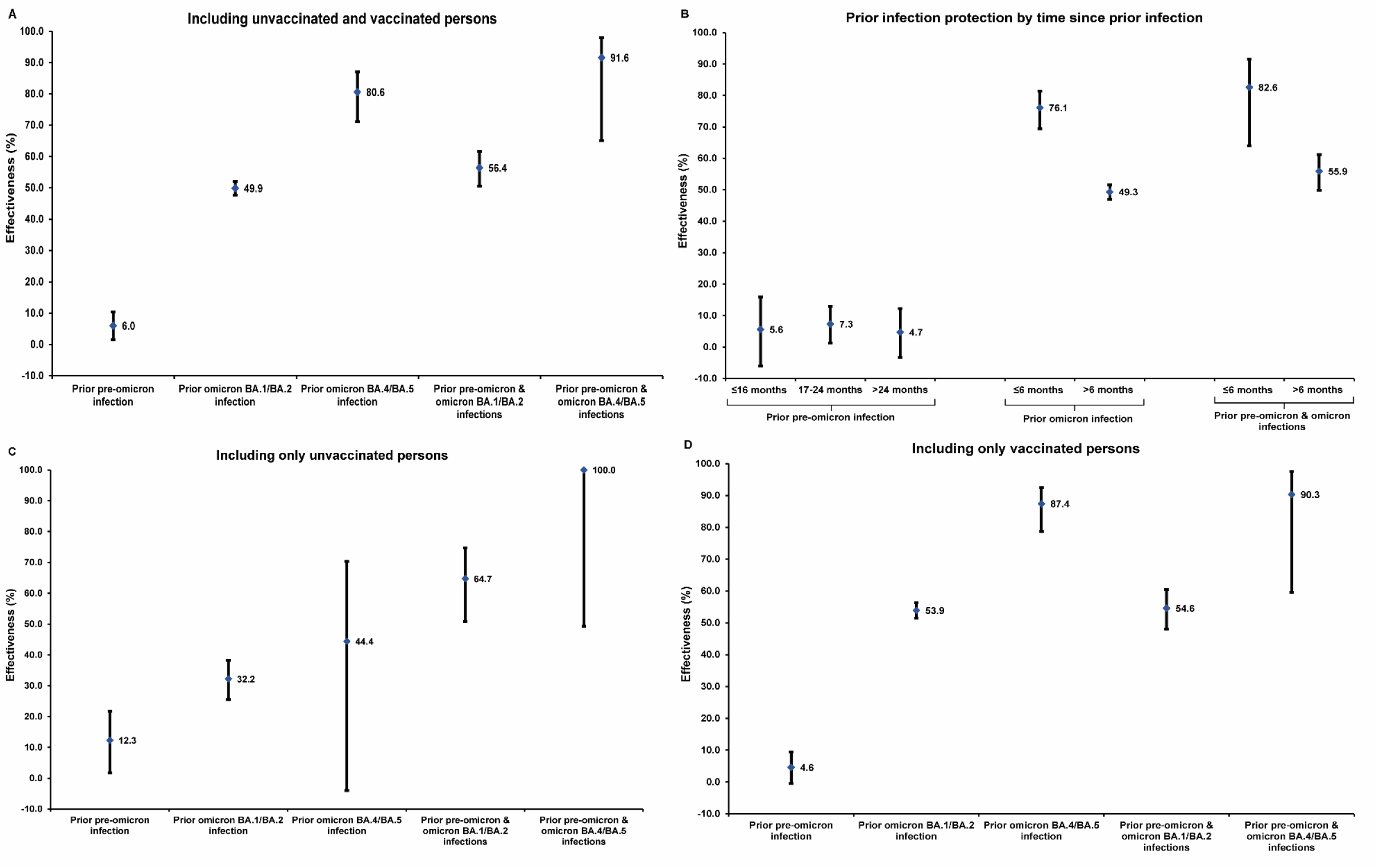
Effectiveness of SARS-CoV-2 prior infection in preventing reinfection, irrespective of symptoms, with the omicron BA.2.75* sublineage in A) both unvaccinated and vaccinated persons, B) both unvaccinated and vaccinated persons stratified by time since prior infection, C) only unvaccinated persons, and D) only vaccinated persons. The study was conducted in Qatar between September 10, 2022 and October 18, 2022.

Similar, but slightly higher protection was observed against symptomatic BA.2.75* reinfection (Table S3). Analyses stratified by time since prior infection indicated waning of protection since prior infection (Figure 1B and Table S4). Analyses stratified by vaccination status indicated that protection from prior infection is higher among those vaccinated, particularly among those who combined index-virus-type vaccination with a prior omicron infection (Figures 1C, 1D, and Table S4). This confirms finding of history of pre-omicron followed by omicron immunity broadening protection against future omicron infection.^4^ Severe COVID-19 was rare (Section S4). Limitations are discussed in Section S2.

Protection against BA.2.75.2 reinfection appears lower than that found against BA.4/BA.5 reinfection.^3^ Protection from prior pre-omicron infection is negligible at this stage of the pandemic, confirming that pre-omicron immunity may not last beyond ∼1 year against omicron infection.^5^ Protection of prior omicron infection was moderate at ∼50% when the prior infection was with BA.1/BA.2, but ∼80% when the prior infection was recent, with BA.4/BA.5, perhaps reflecting progressive immune evasion and gradual natural immunity waning. A combination of pre-omicron and omicron immunity is most protective against BA.2.75* reinfection.

Viral immune evasion may have accelerated recently to overcome high immunity in the global population, thereby also accelerating waning of natural immunity.^5^

## Data Availability

The dataset of this study is a property of the Qatar Ministry of Public Health that was provided to the researchers through a restricted-access agreement that prevents sharing the dataset with a third party or publicly. Future access to this dataset can be considered through a direct application for data access to Her Excellency the Minister of Public Health (https://www.moph.gov.qa/english/Pages/default.aspx). Aggregate data are available within the manuscript and its Supplementary information.

## Oversight

The institutional review boards at Hamad Medical Corporation and Weill Cornell Medicine– Qatar approved this retrospective study with a waiver of informed consent. The study was reported according to the Strengthening the Reporting of Observational Studies in Epidemiology (STROBE) guidelines (Table S5). The authors vouch for the accuracy and completeness of the data and for the fidelity of the study to the protocol. The data set used in this study is the property of the Ministry of Public Health of Qatar and was provided to the researchers through a restricted-access agreement for the preservation of confidentiality of patient data. The funders had no role in the study design; the collection, analysis, or interpretation of the data; or the writing of the manuscript.

### Author contributions

HC co-designed the study, performed the statistical analyses, and co-wrote the first draft of the article. LJA conceived and co-designed the study, led the statistical analyses, and co-wrote the first draft of the article. PT and MRH conducted the multiplex, RT-qPCR variant screening and viral genome sequencing. PVC designed mass PCR testing to allow routine capture of SGTF variants. HY, HAK, and MKS conducted viral genome sequencing. All authors contributed to data collection and acquisition, database development, discussion and interpretation of the results, and to the writing of the manuscript. All authors have read and approved the final manuscript.

## Acknowledgements and support

We acknowledge the many dedicated individuals at Hamad Medical Corporation, the Ministry of Public Health, the Primary Health Care Corporation, the Qatar Biobank, Sidra Medicine, and Weill Cornell Medicine – Qatar for their diligent efforts and contributions to make this study possible.

The authors are grateful for support from the Biomedical Research Program and the Biostatistics, Epidemiology, and Biomathematics Research Core, both at Weill Cornell Medicine-Qatar, as well as for support provided by the Ministry of Public Health, Hamad Medical Corporation, and Sidra Medicine. The authors are also grateful for the Qatar Genome Programme and Qatar University Biomedical Research Center for institutional support for the reagents needed for the viral genome sequencing. Statements made herein are solely the responsibility of the authors. The funders of the study had no role in study design, data collection, data analysis, data interpretation, or writing of the article.

## Supplementary Appendix

### Section S1. Laboratory methods

#### Viral genome sequencing and classification of infections by variant type

Surveillance for the severe acute respiratory syndrome coronavirus 2 (SARS-CoV-2) variants in Qatar is based on viral genome sequencing and multiplex real-time reverse-transcription polymerase chain reaction (RT-qPCR) variant screening^1^ of random positive clinical samples,^2-7^ complemented by deep sequencing of wastewater samples.^4,8,9^ Further details on the viral genome sequencing and multiplex RT-qPCR variant screening throughout the SARS-CoV-2 waves in Qatar can be found in previous publications.^2-7,10-16^

Between September 11, 2022 and October 15, 2022, viral genome sequencing of 587 randomly collected positive samples with polymerase chain reaction (PCR) cycle threshold (Ct) values ≤25 showed that 453 (77.2%) were BA.2.75.X (of which 327, 72.2% were BA.2.75.2), 94 (16.0%) were BA.5.X, 25 (4.3%) were recombinants including XBB, 9 (1.5%) were other BA.2-derived sublineages, 2 (0.3%) were BA.4.X, and 4 (0.7%) could not be classified. Within the 487 BA.2-like subvariants and recombinants, the majority (453, 93.0%) were BA.2.75.X, thus the identification of BA.2-like viruses by RT-qPCR variant screening was used as a proxy for BA.2.75.X.

Between September 10, 2022 and September 25, 2022, RT-qPCR variant screening was performed on 939 random positive clinical samples. In 913 samples, the omicron subvariant could be determined: 695 (76.1%) were BA.2-like, 197 (21.6%) were BA.5-like, 21 (2.3%) were BA.1/BA.4-like subvariants. The RT-qPCR variant screening was validated against virus whole genome sequencing, with a concordance of 98.9% between both methods in a set of 89 random samples including BA.2.X, BA.4.X, and BA.5.X.

#### Real-time reverse-transcription polymerase chain reaction testing

Nasopharyngeal and/or oropharyngeal swabs are collected for PCR testing and placed in Universal Transport Medium (UTM). Aliquots of UTM are: 1) extracted on KingFisher Flex (Thermo Fisher Scientific, USA), MGISP-960 (MGI, China), or ExiPrep 96 Lite (Bioneer, South Korea) followed by testing with RT-qPCR using TaqPath COVID-19 Combo Kits (Thermo Fisher Scientific, USA) on an ABI 7500 FAST (Thermo Fisher Scientific, USA); 2) tested directly on the Cepheid GeneXpert system using the Xpert Xpress SARS-CoV-2 (Cepheid, USA); or 3) loaded directly into a Roche cobas 6800 system and assayed with the cobas SARS-CoV-2 Test (Roche, Switzerland). The first assay targets the viral S, N, and ORF1ab gene regions. The second targets the viral N and E-gene regions, and the third targets the ORF1ab and E-gene regions.

All PCR testing is conducted at the Hamad Medical Corporation Central Laboratory or Sidra Medicine Laboratory, following standardized protocols.

#### Rapid antigen testing

SARS-CoV-2 antigen tests are performed on nasopharyngeal swabs using one of the following lateral flow antigen tests: Panbio COVID-19 Ag Rapid Test Device (Abbott, USA); SARS-CoV-2 Rapid Antigen Test (Roche, Switzerland); Standard Q COVID-19 Antigen Test (SD Biosensor, Korea); or CareStart COVID-19 Antigen Test (Access Bio, USA). All antigen tests are performed at point-of-care according to each manufacturer’s instructions at public or private hospitals and clinics throughout Qatar with prior authorization and training by the Ministry of Public Health (MOPH). Antigen test results are electronically reported to the MOPH in real time using the Antigen Test Management System which is integrated with the national Coronavirus Disease 2019 (COVID-19) database.

### Section S2. Study population, data sources, and study design

This study was conducted in the resident population of Qatar, applying the test-negative, case-control study design^12,13,16,17^ to investigate the protection afforded by prior infection with the severe acute respiratory syndrome coronavirus 2 (SARS-CoV-2) in preventing reinfection with the SARS-CoV-2 BA.2.75* sublineage of the B.1.1.529 (omicron)^18^ variant. Effectiveness of prior infection in preventing reinfection (*PE*_*S*_) was defined as the proportional reduction in susceptibility to infection among those with prior infection versus those without.^12,13,16,17,19^

The study analyzed the national, federated databases for coronavirus disease 2019 (COVID-19) laboratory testing, vaccination, clinical infection data, hospitalization, and death, retrieved from the integrated nationwide digital-health information platform. Databases include all SARS-CoV-2-related data and associated demographic information, with no missing information, since pandemic onset, documenting all polymerase chain reaction (PCR) testing and more recently, rapid antigen (RA) testing conducted at healthcare facilities (from January 5, 2022 onward). Description of laboratory methods for the PCR and RA testing and variant ascertainment are found in Section S1.

Every PCR test (but not every RA test) conducted in Qatar is classified on the basis of symptoms and the reason for testing (clinical symptoms, contact tracing, surveys or random testing campaigns, individual requests, routine healthcare testing, pre-travel, at port of entry, or other). PCR and RA testing is done at a mass scale, where about 5% of the population are tested every week.^7^ Most infections are diagnosed not because of appearance of symptoms, but because of routine testing.^7^ Qatar has unusually young, diverse demographics, in that only 9% of its residents are ≥50 years of age, and 89% are expatriates from over 150 countries.^20,21^ Qatar launched its COVID-19 vaccination program in December of 2020 using the BNT162b2 and mRNA-1273 vaccines.^22^ Further descriptions of the study population and these national databases have been reported previously.^2,3,7,10,21-23^

For estimation of *PE*_*S*_ against BA.2.75* (predominantly BA.2.75.2; Section S1) infection, we exact-matched cases (SARS-CoV-2-positive tests) and controls (SARS-CoV-2-negative tests) in a one-to-five ratio by sex, 10-year age group, nationality, number of coexisting conditions, number of vaccine doses at time of the SARS-CoV-2 test, calendar week of testing, method of testing (PCR or RA), and reason for testing. Matching was done to control for known differences in the risk of exposure to SARS-CoV-2 infection in Qatar.^21,24-27^ Matching by these factors was previously shown to provide adequate control of differences in risk of SARS-CoV-2 exposure in studies of different designs and that included control groups to test for null effects, including test-negative case-control studies.^3,7,22,28,29^

Effectiveness was estimated by comparing odds of prior infection in all cases diagnosed when BA.2.75 dominated incidence (Figure S1 and real-time reverse-transcription (RT-qPCR) variant screening and viral genome sequencing data in Section S1), that is between September 10, 2022 and October 18, 2022 (end of study), to odds of prior infection among controls.

Only the first SARS-CoV-2-positive test during the study period was included for each person (case), while all SARS-CoV-2-negative tests were included for controls. Persons qualified as controls if they had no record of a SARS-CoV-2-positive test during the study. While all PCR tests had a specified reason for testing, only a proportion of RA tests had a specified reason for testing, as some of the testing facilities did not have their electronic system upgraded to capture the reason for testing. Cases and controls were matched by reason for testing, and if reason for testing was not available for the RA test, tests with unspecified reason for testing among cases were matched with tests with unspecified reason for testing among controls.

SARS-CoV-2 reinfection is conventionally defined as a documented infection ≥90 days after an earlier infection, to avoid misclassification of prolonged PCR positivity as reinfection, if a shorter time interval is used.^13,30^ Prior infection was thus defined as a SARS-CoV-2-positive test ≥90 days before this study’s SARS-CoV-2 test. Cases or controls with SARS-CoV-2-positive tests <90 days before the study’s SARS-CoV-2 test were excluded. A symptomatic infection was defined as a SARS-CoV-2 PCR or RA test conducted because of clinical suspicion due to presence of symptoms compatible with a respiratory tract infection.

Every control that met the inclusion criteria and that could be matched to a case was included in the study. Controls were included in the analysis only once. The above inclusion and exclusion criteria were implemented to minimize different types of potential bias, as informed by previous analyses.^7,12,13^

Prior infections were classified as pre-omicron versus omicron based on whether they occurred before or after the omicron wave that started in Qatar on December 19, 2021.^10,12-14,31^ Prior omicron infections were classified by subvariant/sublineage according to when each subvariant/sublineage dominated incidence: BA.1 and BA.2 dominated incidence during December 19, 2021-June 7, 2022,^10,12-14,31^ whereas BA.4 and BA.5 dominated incidence during June 8, 2022-September 9, 2022.^16^

#### Oversight

The Hamad Medical Corporation and Weill Cornell Medicine-Qatar Institutional Review Boards approved this retrospective study with a waiver of informed consent. The study was reported following the Strengthening the Reporting of Observational Studies in Epidemiology (STROBE) guidelines. The STROBE checklist is found in Table S5. The authors vouch for the accuracy and completeness of the data and for the fidelity of the study to the methods protocol. The data set used in this study is the property of the Ministry of Public Health of Qatar and was provided to the researchers through a restricted-access agreement for the preservation of confidentiality of patient data. The funders had no role in the study design; the collection, analysis, or interpretation of the data; or the writing of the manuscript.

#### Statistical analysis

Cases and controls were described using frequency distributions and measures of central tendency and compared using standardized mean differences. A standardized mean difference was defined as the difference in the mean of a covariate between groups, divided by the pooled standard deviation, with values ≤0.1 conventionally indicating adequate balance in matching.^32^ *PE*_*S*_ was derived as one minus the ratio of the odds of prior infection in cases (SARS-CoV-2-positive tests) to the odds of prior infection in controls (SARS-CoV-2-negative tests):^17^

*PE*_*S*_ = 1 – odds ratio of prior infection among cases versus controls.

Odds ratios and associated 95% confidence intervals (CIs) were derived using conditional logistic regression, factoring the matching in the study design. This analytical approach, that also factors matching by number of vaccine doses at time of the SARS-CoV-2 test and calendar week of test, minimizes potential bias due to variation in epidemic phase^33,34^ and roll-out of vaccination during the study.^33,34^ CIs were not adjusted for multiplicity and thus should not be used to infer definitive differences between different groups. Interactions were not investigated.

Additional analyses were conducted. Effectiveness of prior infection against BA.2.75* reinfection was estimated against only symptomatic reinfection. Effectiveness of prior infection against BA.2.75* reinfection was also estimated stratified by time since prior infection and stratified by vaccination status. Statistical analyses were conducted in STATA/SE version 17.0 (Stata Corporation, College Station, TX, USA).

#### Limitations

Ascertainment of the BA.2.75* cases that were included in analysis and of the variant/subvariant status of prior infections was based on calendar time in which each subvariant/sublineage dominated incidence and not based on viral genome sequencing or RT-qPCR variant screening of every infection. However, this approach for ascertainment of variant/subvariant/sublineage status has been shown recently to provide reliable estimates for the protection of prior infection against reinfection with BA.4/BA.5 and is widely used in the literature.^16^ Nevertheless, given overlap in calendar time between the BA.4/BA.5 wave in Qatar^16^ and that of the subsequent BA.2.75* wave, this approach for variant/subvariant/sublineage ascertainment may have led to over-estimation of the protection of prior BA.4/BA.5 infection against reinfection with BA.2.75*, as some of the BA.4/BA.5 infections may have been misclassified as BA.2.75* infections if they occurred during the study, between September 10, 2022 and October 18, 2022. However, the impact of such potential misclassification bias is not expected to be appreciable as the proportion of BA.4/BA.5 infections out of all incident infections was small during the study (Section S1).

With the relatively young population of Qatar, our findings may not be generalizable to other countries where elderly citizens constitute a larger proportion of the total population. With the relatively young population of Qatar,^21,35^ the lower severity of omicron infections,^36-38^ and the time lag between infection and severe forms of COVID-19, there were too small number of confirmed severe,^39^ critical,^39^ and fatal^40^ COVID-19 cases to estimate *PE*_*S*_ against severe forms of COVID-19 due to reinfection.

The study is based on SARS-CoV-2 tests done on individuals currently in Qatar. Qatar has a diverse expatriate population, and it is possible that some persons may have had a prior infection diagnosis while traveling abroad to visit family or for vacation, which would not have been captured in our national databases. However, this is not likely to affect our estimates. It has already been shown that even considerable levels of misclassification of prior infection status had a minimal impact on estimated *PE*_*S*_,^17^ a key strength of the test-negative design.^17^

Misclassification of prior infection status can lead to under-estimation of effectiveness of prior infection in preventing reinfection using the test-negative, case-control study design,^17^ if more than 50% of the population already had a prior infection,^17^ a situation that likely has been reached in Qatar. Nevertheless, this bias did not seem to appreciably affect our earlier analyses for *PE*_*S*_ in the same population of Qatar, as the cohort study designs generated findings that were similar to those of the test-negative, case-control study design.^17,19,23,41-43^ Additionally, we conducted modeling simulations, extending our earlier analyses presented in Ayoub et al,^17^ to investigate the potential effect of this bias.^16^ Assuming *PE*_*S*_ is equal to 40% (instead of 80% in original analysis^17^), a plausible value for pre-omicron protection against omicron subvariants,^12,13,43^ and assuming that 75% of all prior infections are undocumented, the under-estimation of *PE*_*S*_ did not exceed 20 percentage points except when the prevalence of prior infection in the population exceeded 80%, a level that is unlikely to have been reached in Qatar.

Although we used exact matching, the standardized mean differences were larger than zero, and some even slightly larger than 0.1 (Table S1), the conventional limit for adequate matching.^32^ This has occurred because not all cases could be matched to five controls. This, however, should not bias our analyses since cases were exact matched (not matched within a range), and since we used conditional logistic regression which ensures that all comparisons are made within matched sets.

While matching was done for sex, 10-year age group, nationality, number of coexisting conditions, number of vaccine doses at time of the SARS-CoV-2 test, calendar week of testing, method of testing (PCR or RA), and reason for testing, this was not possible for other factors such as geography or occupation, as such data were unavailable. However, Qatar is essentially a city state and infection incidence was broadly distributed across neighborhoods. Nearly 90% of Qatar’s population are expatriates from over 150 countries coming here because of employment;^21^ most are craft and manual workers working in development projects.^21^ Nationality, age, and sex provide a powerful proxy for socio-economic status in this country.^21,24-27^ Nationality alone is strongly associated with occupation.^21,24-27^

It is possible that some people are more likely to be reinfected than others, such as because of occupation or behaviors that involve many unprotected exposures. This may affect the presented estimates. However, matching was done to control for factors known to affect infection exposure in Qatar.^21,24-27^ The matching prescription had already been investigated in previous studies of different epidemiologic designs, and using control groups to test for null effects.^3,7,22,28,29^ These control groups included unvaccinated cohorts versus vaccinated cohorts within two weeks of the first dose,^3,7,28,29^ when vaccine protection is negligible,^44,45^ and mRNA-1273-versus BNT162b2-vaccinated cohorts, also in the first two weeks after the first dose.^22^ These studies have shown that this prescription provides adequate control of the differences in infection exposure.^3,7,22,28,29^ The study was implemented on Qatar’s total population, perhaps thus minimizing the likelihood of bias.

*PE*_*S*_ was assessed using an observational, test-negative, case-control study design,^17^ rather than a cohort study design where individuals are followed up over time. However, the cohort study design applied in earlier analyses to estimate *PE*_*S*_ in the same population of Qatar yielded findings similar to those of the test-negative, case-control study design,^17,19,23,41-43^ supporting the validity of this design in estimating *PE*_*S*_. It even appears that the test-negative study design may be less prone to some forms of bias than the cohort study design.^17^

Nonetheless, one cannot exclude the possibility that in real-world data, bias could arise in unexpected ways, or from unknown sources, such as subtle differences in test-seeking behavior; changes in the pattern of testing due to policy changes, tests’ accessibility, or behavioral differences; or differences in the tendency to get tested between recoverees from prior infection (such as recent recoverees) and those who have not had prior infection or whose prior infection was undocumented. Since SARS-CoV-2 reinfection is conventionally defined as a documented infection ≥90 days after an earlier infection,^13,30^ cases or controls with SARS-CoV-2-positive tests <90 days before the study’s SARS-CoV-2 test were excluded. Therefore, a bias related to testing rates among recent recoverees is not likely to affect our study.

### Section S3. COVID-19 severity, criticality, and fatality classification

Classification of Coronavirus Disease 2019 (COVID-19) case severity (acute-care hospitalizations),^39^ criticality (intensive-care-unit hospitalizations),^39^ and fatality^40^ followed World Health Organization (WHO) guidelines. Assessments were made by trained medical personnel independent of study investigators and using individual chart reviews, as part of a national protocol applied to every hospitalized COVID-19 patient. Each hospitalized COVID-19 patient underwent an infection severity assessment every three days until discharge or death. We classified individuals who progressed to severe, critical, or fatal COVID-19 between the time of the documented infection and the end of the study based on their worst outcome, starting with death,^40^ followed by critical disease,^39^ and then severe disease.^39^

Severe COVID-19 disease was defined per WHO classification as a SARS-CoV-2 infected person with “oxygen saturation of <90% on room air, and/or respiratory rate of >30 breaths/minute in adults and children >5 years old (or ≥60 breaths/minute in children <2 months old or ≥50 breaths/minute in children 2-11 months old or ≥40 breaths/minute in children 1–5 years old), and/or signs of severe respiratory distress (accessory muscle use and inability to complete full sentences, and, in children, very severe chest wall indrawing, grunting, central cyanosis, or presence of any other general danger signs)”.^39^ Detailed WHO criteria for classifying Severe acute respiratory syndrome coronavirus 2 (SARS-CoV-2) infection severity can be found in the WHO technical report.^39^

Critical COVID-19 disease was defined per WHO classification as a SARS-CoV-2 infected person with “acute respiratory distress syndrome, sepsis, septic shock, or other conditions that would normally require the provision of life sustaining therapies such as mechanical ventilation (invasive or non-invasive) or vasopressor therapy”.^39^ Detailed WHO criteria for classifying SARS-CoV-2 infection criticality can be found in the WHO technical report.^39^

COVID-19 death was defined per WHO classification as “a death resulting from a clinically compatible illness, in a probable or confirmed COVID-19 case, unless there is a clear alternative cause of death that cannot be related to COVID-19 disease (e.g. trauma). There should be no period of complete recovery from COVID-19 between illness and death. A death due to COVID-19 may not be attributed to another disease (e.g. cancer) and should be counted independently of preexisting conditions that are suspected of triggering a severe course of COVID-19”. Detailed WHO criteria for classifying COVID-19 death can be found in the WHO technical report.^40^

### Section S4. Severe, critical, or fatal COVID-19 in study population

Of all BA.2.75* cases eligible to be included in the study (29,888 cases; Figure S2), only 6 developed severe forms of coronavirus disease 2019 (COVID-19; Section S3). Specifically, 4 progressed to severe COVID-19,^39^ 1 to critical COVID-19,^39^ and 1 to fatal COVID-19.^40^

Of these, 5 featured in the matched study population (25,449 cases; Figure S2): 3 with severe COVID-19, 1 with critical COVID-19, and 1 with fatal COVID-19. Two of the severe cases and the deceased case had no prior infection, while the rest (one severe case and one critical case) had a prior pre-omicron infection. No severe, critical, or fatal COVID-19 cases were observed among those with a prior omicron infection.

With too few severe, critical, or fatal COVID-19 cases, estimation of effectiveness of prior infection against severe forms of COVID-19 following reinfection with BA.2.75* was not possible.

**Figure S1.**
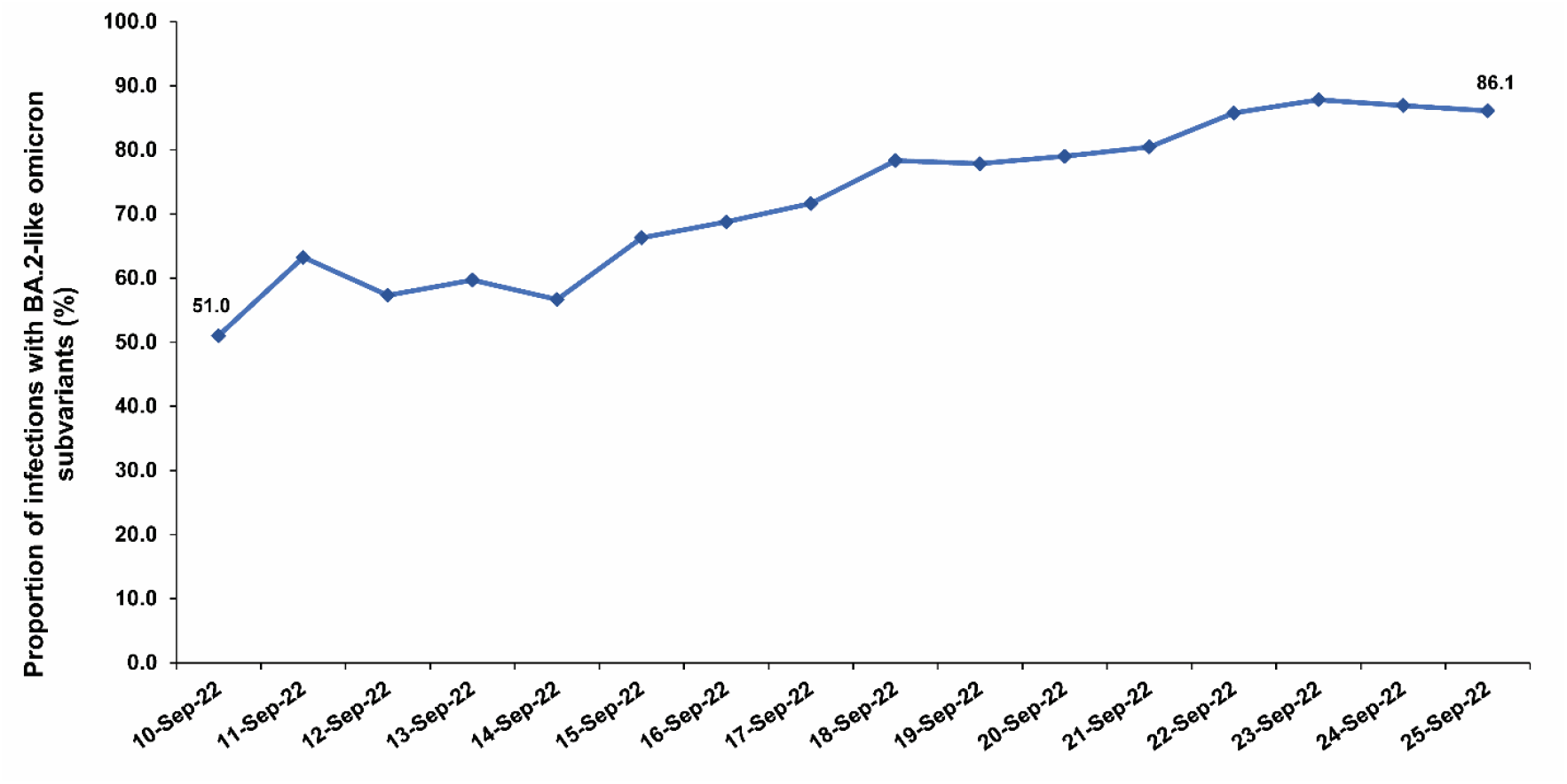
Three-day moving average of the proportion of SARS-CoV-2 infections that are with BA.2-like subvariants in Qatar between September 10, 2022 and September 25, 2022. Subvariant ascertainment was based on multiplex real-time reverse-transcription polymerase chain reaction (RT-qPCR) variant screening^1^ of random positive clinical samples.^2-7^ The identification of BA.2-like subvariants was used as a proxy for BA.2.75* as this was the dominant sublineage within BA.2-like subvariants.

**Figure S2.**
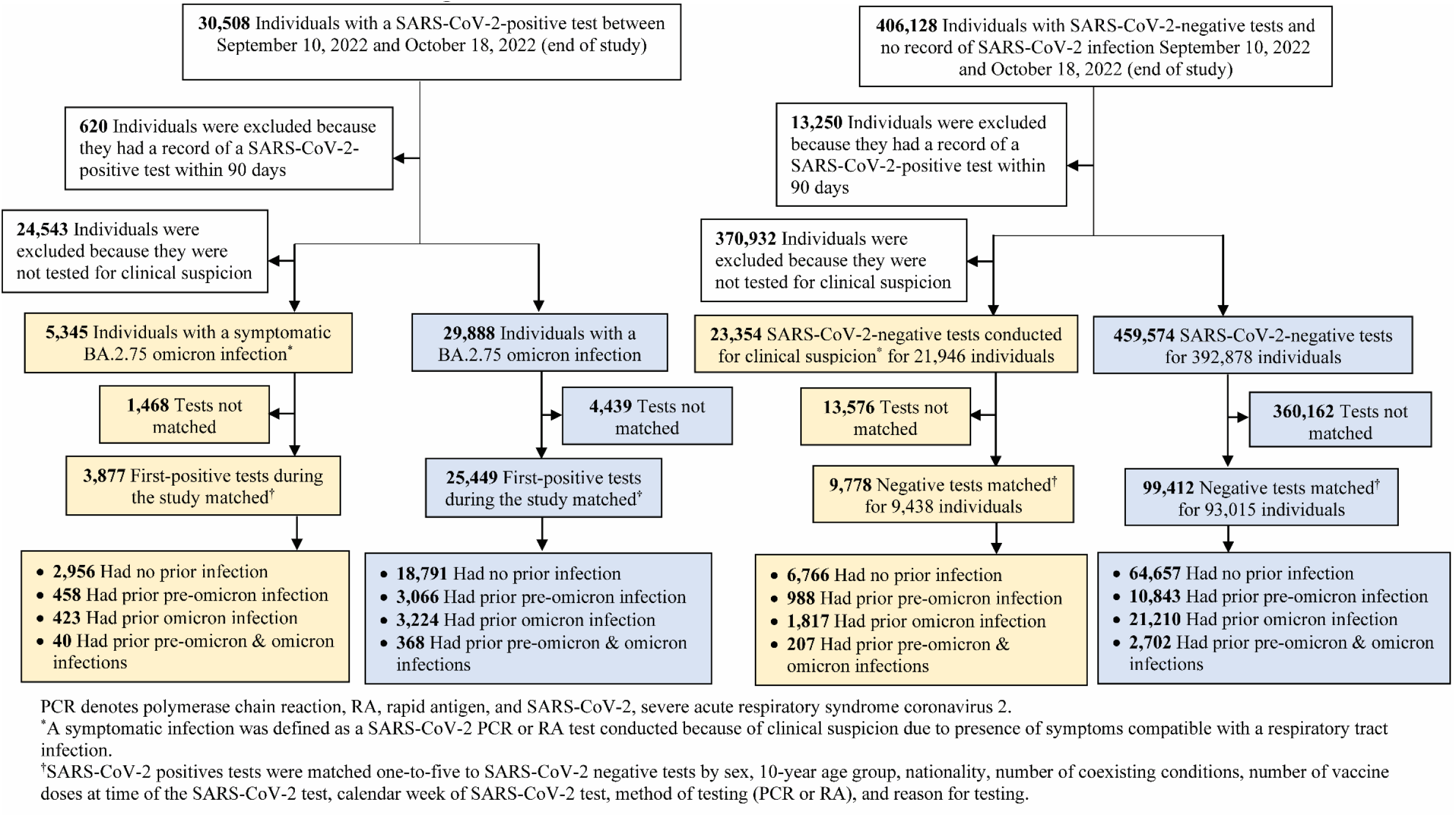
Flowchart describing the population selection process for investigating effectiveness of prior infection in preventing reinfection with the BA.2.75* sublineage.

**Table S1.**
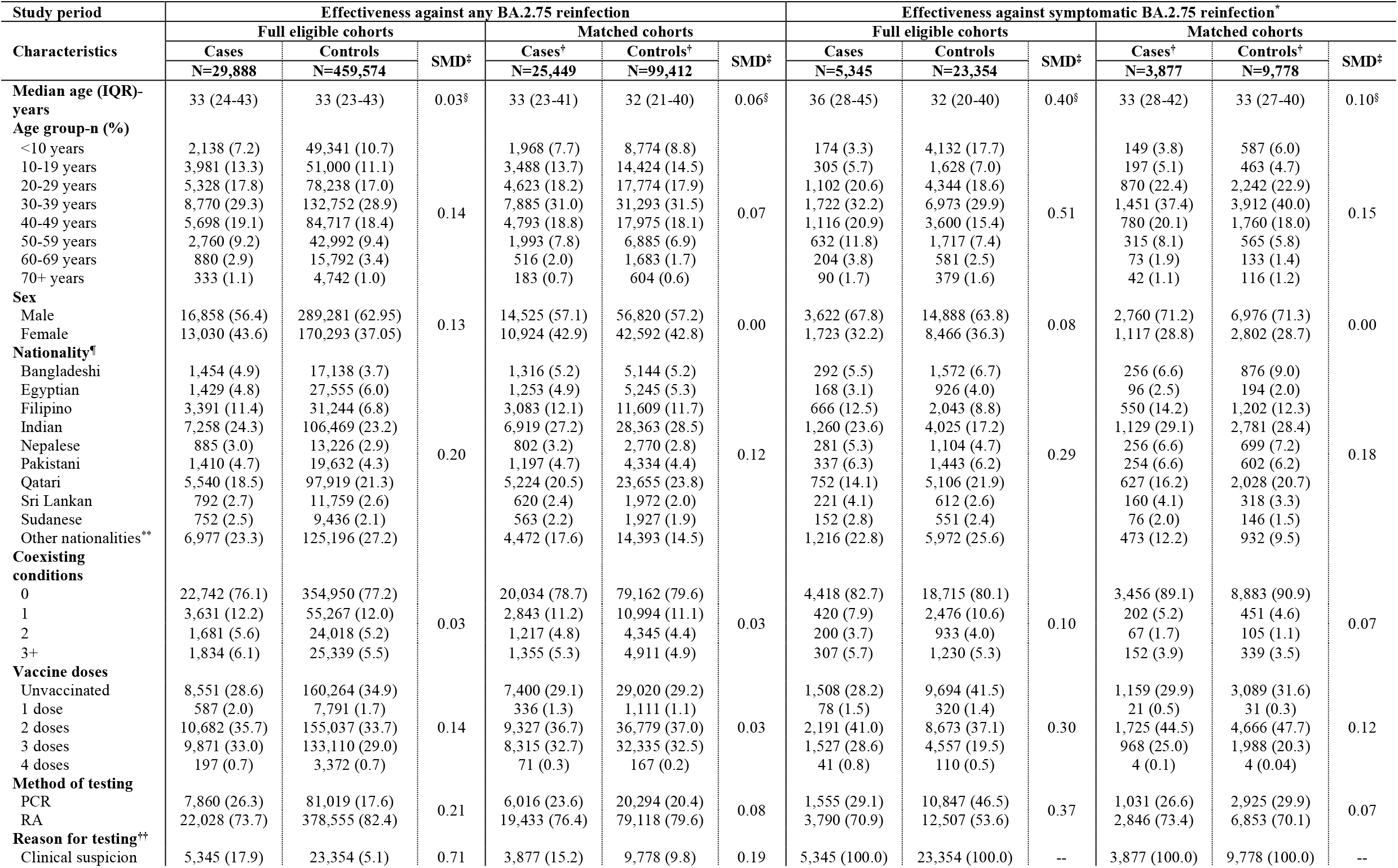
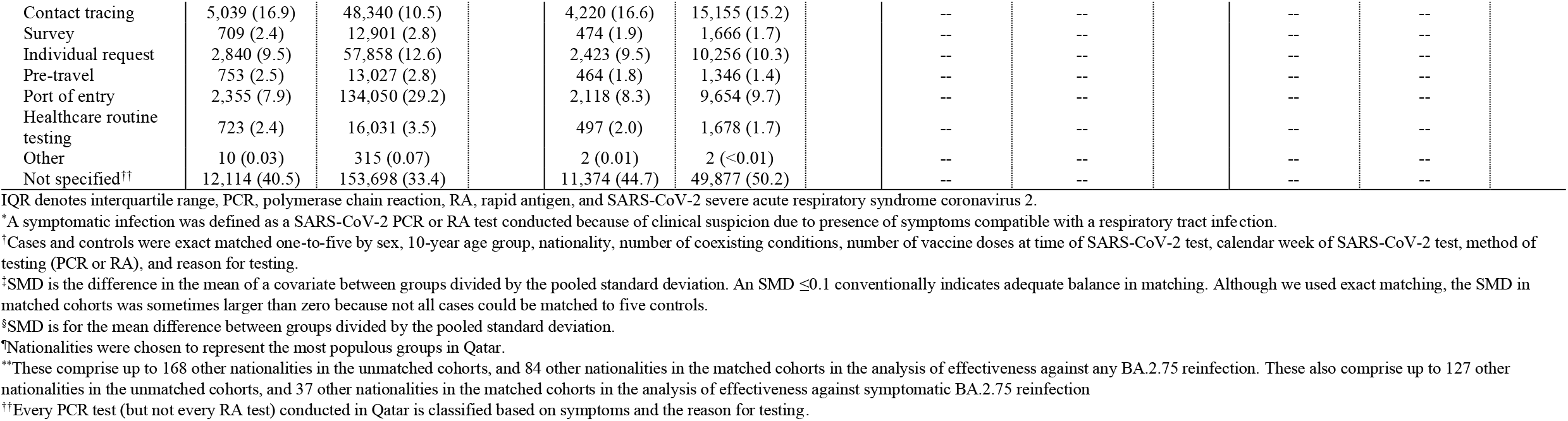
Characteristics of matched cases (SARS-CoV-2-positive tests) and controls (SARS-CoV-2-negative tests) in the analysis assessing effectiveness against any BA.2.75 reinfection regardless of symptoms and in the analysis assessing effectiveness against symptomatic BA.2.75* reinfection. The table is generated for analyses including all SARS-CoV-2 infections diagnosed between September 10, 2022 and October 18, 2022, when BA.2.75* dominated incidence.

| Study period | Effectiveness against any BA.2.75 reinfection |  |  |  |  |  | Effectiveness against symptomatic BA.2.75 reinfection* |  |  |  |  |  |
| --- | --- | --- | --- | --- | --- | --- | --- | --- | --- | --- | --- | --- |
| Characteristics | Full eligible cohorts |  |  | Matched cohorts |  |  | Full eligible cohorts |  |  | Matched cohorts |  |  |
|  | Cases | Controls | SMD<sup>‡</sup> | Cases<sup>†</sup> | Controls<sup>†</sup> | SMD<sup>‡</sup> | Cases | Controls | SMD<sup>‡</sup> | Cases<sup>†</sup> | Controls<sup>†</sup> | SMD<sup>‡</sup> |
|  | N=29,888 | N=459,574 |  | N=25,449 | N=99,412 |  | N=5,345 | N=23,354 |  | N=3,877 | N=9,778 |  |
| **Median age (IQR)-years** | 33 (24-43) | 33 (23-43) | 0.03<sup>§</sup> | 33 (23-41) | 32 (21-40) | 0.06<sup>§</sup> | 36 (28-45) | 32 (20-40) | 0.40<sup>§</sup> | 33 (28-42) | 33 (27-40) | 0.10<sup>§</sup> |
| **Age group-n (%)** |  |  |  |  |  |  |  |  |  |  |  |  |
| <10 years | 2,138 (7.2) | 49,341 (10.7) |  | 1,968 (7.7) | 8,774 (8.8) |  | 174 (3.3) | 4,132 (17.7) |  | 149 (3.8) | 587 (6.0) |  |
| 10-19 years | 3,981 (13.3) | 51,000 (11.1) |  | 3,488 (13.7) | 14,424 (14.5) |  | 305 (5.7) | 1,628 (7.0) |  | 197 (5.1) | 463 (4.7) |  |
| 20-29 years | 5,328 (17.8) | 78,238 (17.0) |  | 4,623 (18.2) | 17,774 (17.9) |  | 1,102 (20.6) | 4,344 (18.6) |  | 870 (22.4) | 2,242 (22.9) |  |
| 30-39 years | 8,770 (29.3) | 132,752 (28.9) |  | 7,885 (31.0) | 31,293 (31.5) |  | 1,722 (32.2) | 6,973 (29.9) |  | 1,451 (37.4) | 3,912 (40.0) |  |
| 40-49 years | 5,698 (19.1) | 84,717 (18.4) | 0.14 | 4,793 (18.8) | 17,975 (18.1) | 0.07 | 1,116 (20.9) | 3,600 (15.4) | 0.51 | 780 (20.1) | 1,760 (18.0) | 0.15 |
| 50-59 years | 2,760 (9.2) | 42,992 (9.4) |  | 1,993 (7.8) | 6,885 (6.9) |  | 632 (11.8) | 1,717 (7.4) |  | 315 (8.1) | 565 (5.8) |  |
| 60-69 years | 880 (2.9) | 15,792 (3.4) |  | 516 (2.0) | 1,683 (1.7) |  | 204 (3.8) | 581 (2.5) |  | 73 (1.9) | 133 (1.4) |  |
| 70+ years | 333 (1.1) | 4,742 (1.0) |  | 183 (0.7) | 604 (0.6) |  | 90 (1.7) | 379 (1.6) |  | 42 (1.1) | 116 (1.2) |  |
| **Sex** |  |  |  |  |  |  |  |  |  |  |  |  |
| Male | 16,858 (56.4) | 289,281 (62.95) |  | 14,525 (57.1) | 56,820 (57.2) |  | 3,622 (67.8) | 14,888 (63.8) |  | 2,760 (71.2) | 6,976 (71.3) |  |
| Female | 13,030 (43.6) | 170,293 (37.05) | 0.13 | 10,924 (42.9) | 42,592 (42.8) | 0.00 | 1,723 (32.2) | 8,466 (36.3) | 0.08 | 1,117 (28.8) | 2,802 (28.7) | 0.00 |
| **Nationality<sup>¶</sup>** |  |  |  |  |  |  |  |  |  |  |  |  |
| Bangladeshi | 1,454 (4.9) | 17,138 (3.7) |  | 1,316 (5.2) | 5,144 (5.2) |  | 292 (5.5) | 1,572 (6.7) |  | 256 (6.6) | 876 (9.0) |  |
| Egyptian | 1,429 (4.8) | 27,555 (6.0) |  | 1,253 (4.9) | 5,245 (5.3) |  | 168 (3.1) | 926 (4.0) |  | 96 (2.5) | 194 (2.0) |  |
| Filipino | 3,391 (11.4) | 31,244 (6.8) |  | 3,083 (12.1) | 11,609 (11.7) |  | 666 (12.5) | 2,043 (8.8) |  | 550 (14.2) | 1,202 (12.3) |  |
| Indian | 7,258 (24.3) | 106,469 (23.2) |  | 6,919 (27.2) | 28,363 (28.5) |  | 1,260 (23.6) | 4,025 (17.2) |  | 1,129 (29.1) | 2,781 (28.4) |  |
| Nepalese | 885 (3.0) | 13,226 (2.9) |  | 802 (3.2) | 2,770 (2.8) |  | 281 (5.3) | 1,104 (4.7) |  | 256 (6.6) | 699 (7.2) |  |
| Pakistani | 1,410 (4.7) | 19,632 (4.3) | 0.20 | 1,197 (4.7) | 4,334 (4.4) | 0.12 | 337 (6.3) | 1,443 (6.2) | 0.29 | 254 (6.6) | 602 (6.2) | 0.18 |
| Qatari | 5,540 (18.5) | 97,919 (21.3) |  | 5,224 (20.5) | 23,655 (23.8) |  | 752 (14.1) | 5,106 (21.9) |  | 627 (16.2) | 2,028 (20.7) |  |
| Sri Lankan | 792 (2.7) | 11,759 (2.6) |  | 620 (2.4) | 1,972 (2.0) |  | 221 (4.1) | 612 (2.6) |  | 160 (4.1) | 318 (3.3) |  |
| Sudanese | 752 (2.5) | 9,436 (2.1) |  | 563 (2.2) | 1,927 (1.9) |  | 152 (2.8) | 551 (2.4) |  | 76 (2.0) | 146 (1.5) |  |
| Other nationalities** | 6,977 (23.3) | 125,196 (27.2) |  | 4,472 (17.6) | 14,393 (14.5) |  | 1,216 (22.8) | 5,972 (25.6) |  | 473 (12.2) | 932 (9.5) |  |
| **Coexisting conditions** |  |  |  |  |  |  |  |  |  |  |  |  |
| 0 | 22,742 (76.1) | 354,950 (77.2) |  | 20,034 (78.7) | 79,162 (79.6) |  | 4,418 (82.7) | 18,715 (80.1) |  | 3,456 (89.1) | 8,883 (90.9) |  |
| 1 | 3,631 (12.2) | 55,267 (12.0) |  | 2,843 (11.2) | 10,994 (11.1) |  | 420 (7.9) | 2,476 (10.6) |  | 202 (5.2) | 451 (4.6) |  |
| 2 | 1,681 (5.6) | 24,018 (5.2) | 0.03 | 1,217 (4.8) | 4,345 (4.4) | 0.03 | 200 (3.7) | 933 (4.0) | 0.10 | 67 (1.7) | 105 (1.1) | 0.07 |
| 3+ | 1,834 (6.1) | 25,339 (5.5) |  | 1,355 (5.3) | 4,911 (4.9) |  | 307 (5.7) | 1,230 (5.3) |  | 152 (3.9) | 339 (3.5) |  |
| **Vaccine doses** |  |  |  |  |  |  |  |  |  |  |  |  |
| Unvaccinated | 8,551 (28.6) | 160,264 (34.9) |  | 7,400 (29.1) | 29,020 (29.2) |  | 1,508 (28.2) | 9,694 (41.5) |  | 1,159 (29.9) | 3,089 (31.6) |  |
| 1 dose | 587 (2.0) | 7,791 (1.7) |  | 336 (1.3) | 1,111 (1.1) |  | 78 (1.5) | 320 (1.4) |  | 21 (0.5) | 31 (0.3) |  |
| 2 doses | 10,682 (35.7) | 155,037 (33.7) | 0.14 | 9,327 (36.7) | 36,779 (37.0) | 0.03 | 2,191 (41.0) | 8,673 (37.1) | 0.30 | 1,725 (44.5) | 4,666 (47.7) | 0.12 |
| 3 doses | 9,871 (33.0) | 133,110 (29.0) |  | 8,315 (32.7) | 32,335 (32.5) |  | 1,527 (28.6) | 4,557 (19.5) |  | 968 (25.0) | 1,988 (20.3) |  |
| 4 doses | 197 (0.7) | 3,372 (0.7) |  | 71 (0.3) | 167 (0.2) |  | 41 (0.8) | 110 (0.5) |  | 4 (0.1) | 4 (0.04) |  |
| **Method of testing** |  |  |  |  |  |  |  |  |  |  |  |  |
| PCR | 7,860 (26.3) | 81,019 (17.6) |  | 6,016 (23.6) | 20,294 (20.4) |  | 1,555 (29.1) | 10,847 (46.5) |  | 1,031 (26.6) | 2,925 (29.9) |  |
| RA | 22,028 (73.7) | 378,555 (82.4) | 0.21 | 19,433 (76.4) | 79,118 (79.6) | 0.08 | 3,790 (70.9) | 12,507 (53.6) | 0.37 | 2,846 (73.4) | 6,853 (70.1) | 0.07 |
| **Reason for testing<sup>††</sup>** |  |  |  |  |  |  |  |  |  |  |  |  |
| Clinical suspicion | 5,345 (17.9) | 23,354 (5.1) | 0.71 | 3,877 (15.2) | 9,778 (9.8) | 0.19 | 5,345 (100.0) | 23,354 (100.0) | -- | 3,877 (100.0) | 9,778 (100.0) | -- |

|  |  |  |  |  |  |  |  |  |  |  |  |
| --- | --- | --- | --- | --- | --- | --- | --- | --- | --- | --- | --- |
| Contact tracing | 5,039 (16.9) | 48,340 (10.5) |  | 4,220 (16.6) | 15,155 (15.2) |  | -- | -- |  | -- | -- |
| Survey | 709 (2.4) | 12,901 (2.8) |  | 474 (1.9) | 1,666 (1.7) |  | -- | -- |  | -- | -- |
| Individual request | 2,840 (9.5) | 57,858 (12.6) |  | 2,423 (9.5) | 10,256 (10.3) |  | -- | -- |  | -- | -- |
| Pre-travel | 753 (2.5) | 13,027 (2.8) |  | 464 (1.8) | 1,346 (1.4) |  | -- | -- |  | -- | -- |
| Port of entry | 2,355 (7.9) | 134,050 (29.2) |  | 2,118 (8.3) | 9,654 (9.7) |  | -- | -- |  | -- | -- |
| Healthcare routine testing | 723 (2.4) | 16,031 (3.5) |  | 497 (2.0) | 1,678 (1.7) |  | -- | -- |  | -- | -- |
| Other | 10 (0.03) | 315 (0.07) |  | 2 (0.01) | 2 (<0.01) |  | -- | -- |  | -- | -- |
| Not specified<sup>††</sup> | 12,114 (40.5) | 153,698 (33.4) |  | 11,374 (44.7) | 49,877 (50.2) |  | -- | -- |  | -- | -- |

IQR denotes interquartile range, PCR, polymerase chain reaction, RA, rapid antigen, and SARS-CoV-2 severe acute respiratory syndrome coronavirus 2.
<sup>\*</sup>A symptomatic infection was defined as a SARS-CoV-2 PCR or RA test conducted because of clinical suspicion due to presence of symptoms compatible with a respiratory tract infection.
<sup>†</sup>Cases and controls were exact matched one-to-five by sex, 10-year age group, nationality, number of coexisting conditions, number of vaccine doses at time of SARS-CoV-2 test, calendar week of SARS-CoV-2 test, method of testing (PCR or RA), and reason for testing.
<sup>‡</sup>SMD is the difference in the mean of a covariate between groups divided by the pooled standard deviation. An SMD $\leq 0.1$ conventionally indicates adequate balance in matching. Although we used exact matching, the SMD in matched cohorts was sometimes larger than zero because not all cases could be matched to five controls.
<sup>§</sup>SMD is for the mean difference between groups divided by the pooled standard deviation.
<sup>\*</sup>Nationalities were chosen to represent the most populous groups in Qatar.
<sup>\*\*</sup>These comprise up to 168 other nationalities in the unmatched cohorts, and 84 other nationalities in the matched cohorts in the analysis of effectiveness against any BA.2.75 reinfection. These also comprise up to 127 other nationalities in the unmatched cohorts, and 37 other nationalities in the matched cohorts in the analysis of effectiveness against symptomatic BA.2.75 reinfection
<sup>††</sup>Every PCR test (but not every RA test) conducted in Qatar is classified based on symptoms and the reason for testing.

**Table S2.** Representativeness of study participants.

| Category |  |
| --- | --- |
| Disease, problem, or condition under investigation | Effectiveness of prior infection in preventing reinfection with SARS-CoV-2 BA.2.75* sublineage of omicron subvariant BA.2. |
| Special considerations related to |  |
| Sex and gender | The effectiveness estimates were derived by comparing cases (SARS-CoV-2-positive tests) and controls (SARS-CoV-2-negative tests) with respect to prior infection. Cases and controls were exact-matched by sex to control for potential differences in the risk of exposure to SARS-CoV-2 infection by sex. |
| Age | Cases and controls were exact-matched by 10-year age group to control for potential differences in the risk of exposure to SARS-CoV-2 infection by age. Nonetheless, with the young population of Qatar, our findings may not be generalizable to other countries where elderly citizens constitute a larger proportion of the total population. |
| Race or ethnicity group | Cases and controls were exact-matched by nationality to control for potential differences in the risk of exposure to SARS-CoV-2 infection by nationality. Nationality is associated with race and ethnicity in the population of Qatar. |
| Geography | Individual-level data on geography were not available, but Qatar is essentially a city state and infection incidence was broadly distributed across the country's neighborhoods/areas. Cohorts were exact-matched by nationality to control for potential differences in the risk of exposure to SARS-CoV-2 infection by nationality. Qatar has unusually diverse demographics in that 89% of the population are international expatriate residents coming from over 150 countries from all world regions. |
| Other considerations | Individual-level data on occupation were not available but matching by nationality may have (partially) controlled the differences in occupational risk, in consideration of the association between nationality and occupation in Qatar. |
| Overall representativeness of this study | The study was based on the total population of Qatar and thus the study population is broadly representative of the diverse, by national background, but young and predominantly male, total population of Qatar. While there could be differences in the risk of exposure to SARS-CoV-2 infection by sex, age, nationality, number of coexisting conditions, number of vaccine doses at time of SARS-CoV-2 test, calendar week of SARS-CoV-2 test, method of testing and reason for testing, cohorts were exact-matched by these factors to control for their potential impact on our estimates. Given that only 9% of the population of Qatar are $\geq 50$ years of age, our estimates may not be generalizable to other countries where elderly citizens constitute a larger proportion of the total population. |

SARS-CoV-2 denotes severe acute respiratory syndrome coronavirus 2.

**Table S3.**
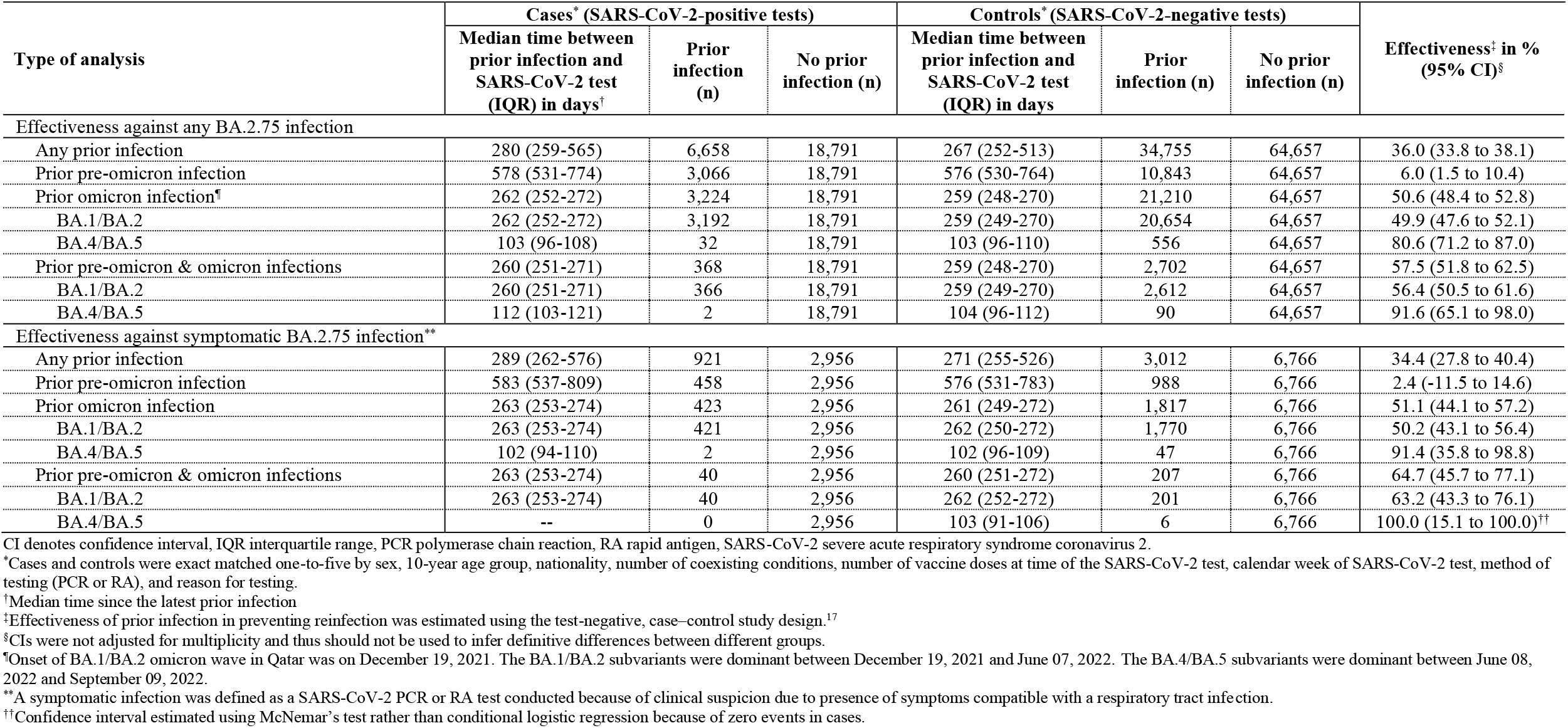
Effectiveness of previous SARS-CoV-2 infection in preventing reinfection with the omicron BA.2.75* sublineage using all SARS-CoV-2 infections diagnosed between September 10, 2022 and October 18, 2022, when BA.2.75* dominated incidence.

**Table S4.** Sensitivity analyses. Effectiveness of SARS-CoV-2 prior infection in preventing any reinfection with the omicron BA.2.75* sublineage using all SARS-CoV-2 infections diagnosed between September 10, 2022 and October 18, 2022, when BA.2.75* dominated incidence, after A) including only unvaccinated individuals, B) including only vaccinated individuals, and C) stratifying by time since prior infection.

| Type of analysis | Cases* (SARS-CoV-2-positive tests) |  |  | Controls* (SARS-CoV-2-negative tests) |  |  | Effectiveness† in %<br>(95% CI)‡ |
| --- | --- | --- | --- | --- | --- | --- | --- |
|  | Median time between prior infection and SARS-CoV-2 test (IQR) in days† | Prior infection (n) | No prior infection (n) | Median time between prior infection and SARS-CoV-2 test (IQR) in days† | Prior infection (n) | No prior infection (n) |  |
| Effectiveness against any BA.2.75 infection |  |  |  |  |  |  |  |
| A) Sensitivity analysis—Including only unvaccinated individuals |  |  |  |  |  |  |  |
| Any prior infection | 270 (253-489) | 1,267 | 6,133 | 265 (249-384) | 6,437 | 22,583 | 27.9 (22.3 to 33.1) |
| Prior pre-omicron infection | 540 (398-614) | 477 | 6,133 | 542 (423-682) | 1,999 | 22,583 | 12.3 (1.7 to 21.7) |
| Prior omicron infection¶ | 258 (246-268) | 741 | 6,133 | 256 (244-267) | 3,933 | 22,583 | 32.5 (25.8 to 38.5) |
| BA.1/BA.2 | 258 (246-269) | 728 | 6,133 | 256 (245-267) | 3,829 | 22,583 | 32.2 (25.5 to 38.3) |
| BA.4/BA.5 | 97 (92-103) | 13 | 6,133 | 101 (95-110) | 104 | 22,583 | 44.4 (-4.0 to 70.3) |
| Prior pre-omicron & omicron infections | 253 (245-265) | 49 | 6,133 | 257 (245-268) | 505 | 22,583 | 65.5 (51.9 to 75.2) |
| BA.1/BA.2 | 253 (245-265) | 49 | 6,133 | 257 (245-268) | 496 | 22,583 | 64.7 (50.8 to 74.7) |
| BA.4/BA.5 | -- | 0 | 6,133 | 107 (107-114) | 9 | 22,583 | 100.0 (49.3 to 100.0)** |
| B) Sensitivity analysis—Including only vaccinated individuals |  |  |  |  |  |  |  |
| Any prior infection | 284 (261-580) | 5,391 | 12,658 | 268 (253-525) | 28,318 | 42,074 | 37.9 (35.5 to 40.2) |
| Prior pre-omicron infection | 588 (537-789) | 2,589 | 12,658 | 586 (537-777) | 8,844 | 42,074 | 4.6 (-0.5 to 9.5) |
| Prior omicron infection¶ | 263 (253-273) | 2,483 | 12,658 | 260 (249-270) | 17,277 | 42,074 | 54.8 (52.4 to 57.1) |
| BA.1/BA.2 | 263 (254-273) | 2,464 | 12,658 | 260 (250-271) | 16,825 | 42,074 | 53.9 (51.5 to 56.3) |
| BA.4/BA.5 | 105 (97-108) | 19 | 12,658 | 103 (96-110) | 452 | 42,074 | 87.4 (78.7 to 92.5) |
| Prior pre-omicron & omicron infections | 261 (252-271) | 319 | 12,658 | 259 (248-270) | 2,197 | 42,074 | 55.9 (49.4 to 61.5) |
| BA.1/BA.2 | 261 (252-271) | 317 | 12,658 | 259 (249-270) | 2,116 | 42,074 | 54.6 (48.0 to 60.4) |
| BA.4/BA.5 | 112 (103-121) | 2 | 12,658 | 103 (95-111) | 81 | 42,074 | 90.3 (59.6 to 97.6) |
| C) Sensitivity analysis—Stratifying by time since the latest prior infection |  |  |  |  |  |  |  |
| Any prior infection |  |  |  |  |  |  |  |
| ≤8 months | 227 (191-236) | 411 | 18,791 | 222 (140-235) | 4,035 | 64,657 | 66.9 (62.9 to 70.4) |
| 9-16 months | 266 (256-277) | 3,667 | 18,791 | 263 (253-274) | 21,596 | 64,657 | 44.9 (42.4 to 47.2) |
| >16 months | 610 (548-799) | 2,580 | 18,791 | 607 (547-785) | 9,124 | 64,657 | 6.2 (1.2 to 10.8) |
| Prior pre-omicron infection |  |  |  |  |  |  |  |
| ≤16 months | 359 (305-420) | 479 | 18,791 | 370 (313-414) | 1,701 | 64,657 | 5.6 (-6.0 to 15.9) |
| 17-24 months | 558 (537-595) | 1,591 | 18,791 | 558 (536-595) | 5,721 | 64,657 | 7.3 (1.2 to 13.0) |
| >24 months | 826 (780-853) | 996 | 18,791 | 818 (772-853) | 3,421 | 64,657 | 4.7 (-3.3 to 12.2) |
| Prior omicron infection |  |  |  |  |  |  |  |
| ≤6 months | 133 (105-162) | 82 | 18,791 | 115 (102-137) | 1,122 | 64,657 | 76.1 (69.4 to 81.4) |
| >6 months | 263 (253-272) | 3,142 | 18,791 | 260 (250-270) | 20,088 | 64,657 | 49.3 (46.9 to 51.5) |
| Prior pre-omicron & omicron infections |  |  |  |  |  |  |  |
| ≤6 months | 141 (126-145) | 9 | 18,791 | 114 (101-133) | 165 | 64,657 | 82.6 (64.0 to 91.6) |
| >6 months | 260 (252-271) | 359 | 18,791 | 260 (250-270) | 2,537 | 64,657 | 55.9 (49.9 to 61.2) |

CI denotes confidence interval, IQR interquartile range, PCR polymerase chain reaction, RA rapid antigen, and SARS-CoV-2 severe acute respiratory syndrome coronavirus 2.
\*Cases and controls were exact matched one-to-five by sex, 10-year age group, nationality, number of coexisting conditions, number of vaccine doses at time of the SARS-CoV-2 test, calendar week of SARS-CoV-2 test, method of testing (PCR or RA), and reason for testing.
<sup>7</sup>Median time since the latest prior infection
<sup>8</sup>Effectiveness of prior infection in preventing reinfection was estimated using the test-negative, case-control study design.<sup>17</sup>
<sup>8</sup>CIs were not adjusted for multiplicity and thus should not be used to infer definitive differences between different groups.
<sup>9</sup>Onset of BA.1/BA.2 omicron wave in Qatar was on December 19, 2021. The BA.1/BA.2 subvariants were dominant between December 19, 2021 and June 07, 2022. The BA.4/BA.5 subvariants were dominant between June 08, 2022 and September 09, 2022.
<sup>10</sup>Confidence interval estimated using McNemar's test rather than conditional logistic regression because of zero events in cases.

**Table S5.**
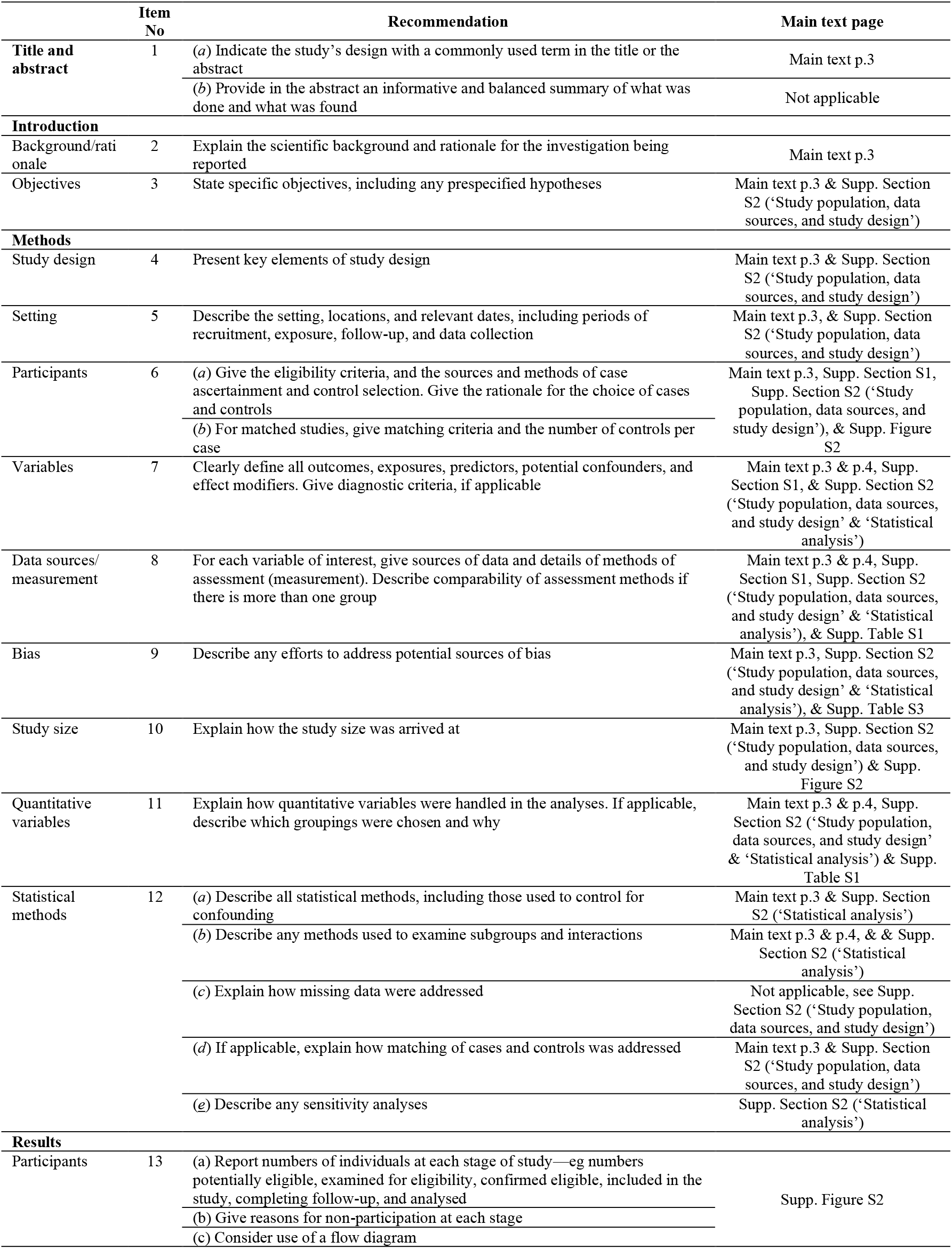
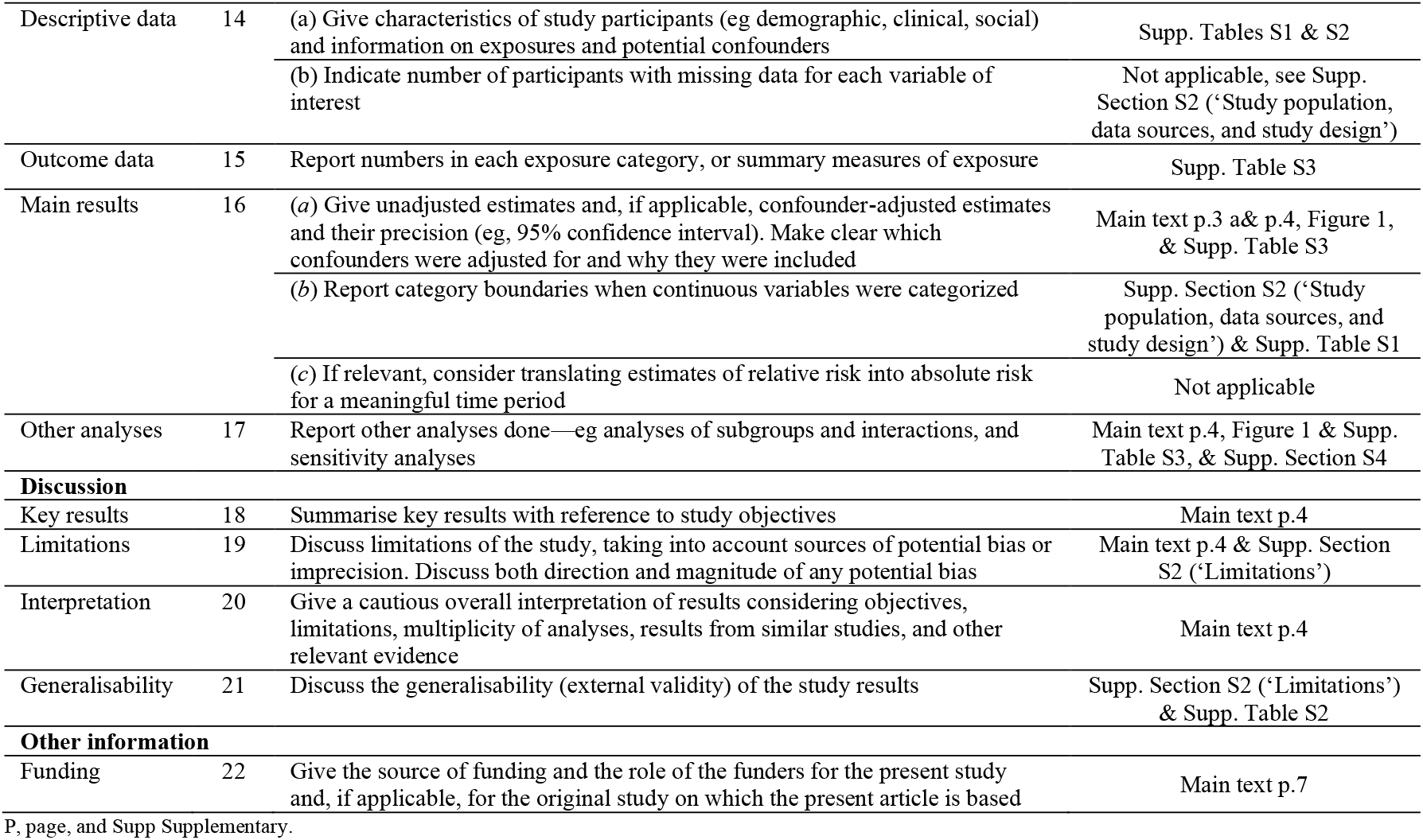
STROBE checklist for case-control studies.

## References

1. Sheward DJ, Kim C, Fischbach J, et al. Omicron sublineage BA.2.75.2 exhibits extensive escape from neutralising antibodies. Lancet Infect Dis 2022.

2. Ayoub HH, Tomy M, Chemaitelly H, et al. Estimating protection afforded by prior infection in preventing reinfection: Applying the test-negative study design. medRxiv 2022:2022.01.02.22268622.

3. Altarawneh HN, Chemaitelly H, Ayoub HH, et al. Protective Effect of Previous SARS-CoV-2 Infection against Omicron BA.4 and BA.5 Subvariants. New England Journal of Medicine 2022.

4. Chemaitelly H, Ayoub HH, Tang P, et al. Immune Imprinting and Protection against Repeat Reinfection with SARS-CoV-2. N Engl J Med 2022.

5. Chemaitelly H, Nagelkerke N, Ayoub HH, et al. Duration of immune protection of SARS-CoV-2 natural infection against reinfection. J Travel Med 2022.

## References

1. Multiplexed RT-qPCR to screen for SARS-COV-2 B.1.1.7, B.1.351, and P.1 variants of concern V.3. dx.doi.org/10.17504/protocols.io.br9vm966. 2021. (Accessed June 6, 2021, at https://www.protocols.io/view/multiplexed-rt-qpcr-to-screen-for-sars-cov-2-b-1-1-br9vm966.)

2. Abu-Raddad LJ, Chemaitelly H, Butt AA, National Study Group for Covid Vaccination. Effectiveness of the BNT162b2 Covid-19 Vaccine against the B.1.1.7 and B.1.351 Variants. N Engl J Med 2021;385:187–9.

3. Chemaitelly H, Yassine HM, Benslimane FM, et al. mRNA-1273 COVID-19 vaccine effectiveness against the B.1.1.7 and B.1.351 variants and severe COVID-19 disease in Qatar. Nat Med 2021;27:1614–21.

4. Qatar viral genome sequencing data. Data on randomly collected samples. https://www.gisaid.org/phylodynamics/global/nextstrain/. 2021. at https://www.gisaid.org/phylodynamics/global/nextstrain/.)

5. Benslimane FM, Al Khatib HA, Al-Jamal O, et al. One Year of SARS-CoV-2: Genomic Characterization of COVID-19 Outbreak in Qatar. Front Cell Infect Microbiol 2021;11:768883.

6. Hasan MR, Kalikiri MKR, Mirza F, et al. Real-Time SARS-CoV-2 Genotyping by High-Throughput Multiplex PCR Reveals the Epidemiology of the Variants of Concern in Qatar. Int J Infect Dis 2021;112:52–4.

7. Chemaitelly H, Tang P, Hasan MR, et al. Waning of BNT162b2 Vaccine Protection against SARS-CoV-2 Infection in Qatar. N Engl J Med 2021;385:e83.

8. Saththasivam J, El-Malah SS, Gomez TA, et al. COVID-19 (SARS-CoV-2) outbreak monitoring using wastewater-based epidemiology in Qatar. Sci Total Environ 2021;774:145608.

9. El-Malah SS, Saththasivam J, Jabbar KA, et al. Application of human RNase P normalization for the realistic estimation of SARS-CoV-2 viral load in wastewater: A perspective from Qatar wastewater surveillance. Environ Technol Innov 2022;27:102775.

10. Abu-Raddad LJ, Chemaitelly H, Ayoub HH, et al. Effect of mRNA Vaccine Boosters against SARS-CoV-2 Omicron Infection in Qatar. N Engl J Med 2022;386:1804–16.

11. Tang P, Hasan MR, Chemaitelly H, et al. BNT162b2 and mRNA-1273 COVID-19 vaccine effectiveness against the SARS-CoV-2 Delta variant in Qatar. Nat Med 2021;27:2136–43.

12. Altarawneh HN, Chemaitelly H, Ayoub HH, et al. Effects of Previous Infection and Vaccination on Symptomatic Omicron Infections. N Engl J Med 2022;387:21–34.

13. Altarawneh HN, Chemaitelly H, Hasan MR, et al. Protection against the Omicron Variant from Previous SARS-CoV-2 Infection. N Engl J Med 2022;386:1288–90.

14. Chemaitelly H, Ayoub HH, AlMukdad S, et al. Duration of mRNA vaccine protection against SARS-CoV-2 Omicron BA.1 and BA.2 subvariants in Qatar. Nat Commun 2022;13:3082.

15. Qassim SH, Chemaitelly H, Ayoub HH, et al. Effects of BA.1/BA.2 subvariant, vaccination, and prior infection on infectiousness of SARS-CoV-2 omicron infections. J Travel Med 2022.

16. Altarawneh HN, Chemaitelly H, Ayoub HH, et al. Protective Effect of Previous SARS-CoV-2 Infection against Omicron BA.4 and BA.5 Subvariants. N Engl J Med 2022.

17. Ayoub HH, Tomy M, Chemaitelly H, et al. Estimating protection afforded by prior infection in preventing reinfection: Applying the test-negative study design. medRxiv 2022:2022.01.02.22268622.

18. World Health Organization. Tracking SARS-CoV-2 variants. Available from: https://www.who.int/en/activities/tracking-SARS-CoV-2-variants/. 2021.

19. Abu-Raddad LJ, Chemaitelly H, Coyle P, et al. SARS-CoV-2 antibody-positivity protects against reinfection for at least seven months with 95% efficacy. EClinicalMedicine 2021;35:100861.

20. Planning and Statistics Authority-State of Qatar. Qatar Monthly Statistics. Available from: https://www.psa.gov.qa/en/pages/default.aspx. Accessed on: May 26, 2020. 2020.

21. Abu-Raddad LJ, Chemaitelly H, Ayoub HH, et al. Characterizing the Qatar advanced-phase SARS-CoV-2 epidemic. Sci Rep 2021;11:6233.

22. Abu-Raddad LJ, Chemaitelly H, Bertollini R, National Study Group for Covid Vaccination. Effectiveness of mRNA-1273 and BNT162b2 Vaccines in Qatar. N Engl J Med 2022;386:799–800.

23. Chemaitelly H, Bertollini R, Abu-Raddad LJ, National Study Group for Covid Epidemiology. Efficacy of Natural Immunity against SARS-CoV-2 Reinfection with the Beta Variant. N Engl J Med 2021;385:2585–6.

24. Ayoub HH, Chemaitelly H, Seedat S, et al. Mathematical modeling of the SARS-CoV-2 epidemic in Qatar and its impact on the national response to COVID-19. J Glob Health 2021;11:05005.

25. Coyle PV, Chemaitelly H, Ben Hadj Kacem MA, et al. SARS-CoV-2 seroprevalence in the urban population of Qatar: An analysis of antibody testing on a sample of 112,941 individuals. iScience 2021;24:102646.

26. Al-Thani MH, Farag E, Bertollini R, et al. SARS-CoV-2 Infection Is at Herd Immunity in the Majority Segment of the Population of Qatar. Open Forum Infect Dis 2021;8:ofab221.

27. Jeremijenko A, Chemaitelly H, Ayoub HH, et al. Herd Immunity against Severe Acute Respiratory Syndrome Coronavirus 2 Infection in 10 Communities, Qatar. Emerg Infect Dis 2021;27:1343–52.

28. Abu-Raddad LJ, Chemaitelly H, Yassine HM, et al. Pfizer-BioNTech mRNA BNT162b2 Covid-19 vaccine protection against variants of concern after one versus two doses. J Travel Med 2021;28.

29. Abu-Raddad LJ, Chemaitelly H, Bertollini R, National Study Group for Covid Vaccination. Waning mRNA-1273 Vaccine Effectiveness against SARS-CoV-2 Infection in Qatar. N Engl J Med 2022;386:1091–3.

30. Pilz S, Theiler-Schwetz V, Trummer C, Krause R, Ioannidis JPA. SARS-CoV-2 reinfections: Overview of efficacy and duration of natural and hybrid immunity. Environ Res 2022:112911.

31. Chemaitelly H, Ayoub HH, Coyle P, et al. Protection of Omicron sub-lineage infection against reinfection with another Omicron sub-lineage. Nat Commun 2022;13:4675.

32. Austin PC. Using the Standardized Difference to Compare the Prevalence of a Binary Variable Between Two Groups in Observational Research. Communications in Statistics - Simulation and Computation 2009;38:1228–34.

33. Jackson ML, Nelson JC. The test-negative design for estimating influenza vaccine effectiveness. Vaccine 2013;31:2165–8.

34. Jacoby P, Kelly H. Is it necessary to adjust for calendar time in a test negative design?: Responding to:

35. Jackson ML, Nelson JC. The test negative design for estimating influenza vaccine effectiveness. Vaccine 2013;31(April (17)):2165-8. Vaccine 2014;32:2942.

36. Seedat S, Chemaitelly H, Ayoub HH, et al. SARS-CoV-2 infection hospitalization, severity, criticality, and fatality rates in Qatar. Sci Rep 2021;11:18182.

37. Butt AA, Dargham SR, Loka S, et al. COVID-19 Disease Severity in Children Infected with the Omicron Variant. Clin Infect Dis 2022.

38. Butt AA, Dargham SR, Coyle P, et al. COVID-19 Disease Severity in Persons Infected With Omicron BA.1 and BA.2 Sublineages and Association With Vaccination Status. JAMA Intern Med 2022.

39. World Health Organization. COVID-19 clinical management: living guidance. Available from: https://www.who.int/publications/i/item/WHO-2019-nCoV-clinical-2021-1. Accessed on: May 15, 2021. 2021.

40. World Health Organization. International guidelines for certification and classification (coding) of COVID-19 as cause of death. Available from: https://www.who.int/classifications/icd/Guidelines_Cause_of_Death_COVID-19-20200420-EN.pdf?ua=1. Document Number: WHO/HQ/DDI/DNA/CAT. Accessed on May 15, 2021. 2020.

41. Abu-Raddad LJ, Chemaitelly H, Ayoub HH, et al. Introduction and expansion of the SARS-CoV-2 B.1.1.7 variant and reinfections in Qatar: A nationally representative cohort study. PLoS Med 2021;18:e1003879.

42. Abu-Raddad LJ, Chemaitelly H, Malek JA, et al. Assessment of the Risk of Severe Acute Respiratory Syndrome Coronavirus 2 (SARS-CoV-2) Reinfection in an Intense Reexposure Setting. Clin Infect Dis 2021;73:e1830–e40.

43. Chemaitelly H, Nagelkerke N, Ayoub HH, et al. Duration of immune protection of SARS-CoV-2 natural infection against reinfection. J Travel Med 2022.

44. Polack FP, Thomas SJ, Kitchin N, et al. Safety and Efficacy of the BNT162b2 mRNA Covid-19 Vaccine. N Engl J Med 2020;383:2603–15.

45. Baden LR, El Sahly HM, Essink B, et al. Efficacy and Safety of the mRNA-1273 SARS-CoV-2 Vaccine. N Engl J Med 2021;384:403–16.

